# COVID-19, a social disease in Paris: a socio-economic wide association study on hospitalized patients highlights low-income neighbourhood as a key determinant of severe COVID-19 incidence during the first wave of the epidemic

**DOI:** 10.1101/2020.10.30.20222901

**Authors:** Anne-Sophie Jannot, Hector Coutouris, Anita Burgun, Sandrine Katsahian, Bastien Rance, on behalf of the AP-HP / Universities / Inserm COVID-19 research collaboration

**Affiliations:** Université de Paris, F-75015 Paris, France; Assistance Publique-Hôpitaux de Paris (AP-HP), Service d’informatique médicale, biostatistiques et santé publique, Hôpital Européen Georges Pompidou, Paris, France; Centre de Recherche des Cordeliers, Sorbonne Université, Inserm, Université de Paris, f-75006, Paris, France, Paris, France

**Keywords:** clinical datawarehouse, ecological study, socio-economic risk factors, machine learning

## Abstract

**Background:** Studies have already shown that many environmental factors are associated with COVID-19 incidence. However, none have studied a very large set of socio-economic indicators and analysed to what extent these factors could highlight populations at high risk for COVID-19. We propose here a new approach, a socio-economic wide study, to pinpoint subgroups with a high incidence of COVID-19, and illustrated this approach using hospitalized cases in Paris area.

**Methods:** We extracted 303 socio-economic indicators from French census data for the 855 residential units in Paris and assessed their association with COVID-19 hospitalization risk. We then fitted a predictive model using a penalized regression on these indicators to predict the incidence of patient hospitalization for COVID-19 in Paris.

**Findings:** The most associated indicator was income, corresponding to the 3^rd^ decile of the population (OR= 0.11, CI95% [0.05; 0.20]). A model including only income achieves a high performance in both the training set (AUC=0.78, CI95%: 0.72-0.85) and the test set (AUC=0.79 (CI95%: 0.71-0.87). Overall, the 45% most deprived areas gathered 86% of the areas with a high incidence of COVID-19 hospitalized cases.

**Interpretation:** During the first wave of the epidemic, income predicted Paris areas at risk for a high incidence of patients hospitalized for COVID-19 with a high performance. Socio-economic wide association studies, collecting passively data from hospitalized cases, therefore not necessitating any effort for health caregivers, is of particular interest in such a period of hospital overcrowding as it provides real-time indirect information on populations having high COVID-19 incidence.

## Introduction

The COVID-19 outbreak has been raging worldwide since late 2019, killing thousands and disrupting health systems with hundreds of hospitalized patients. In France, the peak number of positively tested patients was reached on the 31st of March, with 7578 new cases, most of which were located in the eastern part of France and the greater Paris area. Many environmental factors may contribute to the spread of the outbreak, including population density, pollution, demographics, education and income (1). Paris, the capital of France, is the largest city in the country, with 2 187 526 inhabitants. Its density is one of the highest in the world, with 20,754 inhabitants per squared kilometre. By comparison, New York City, NY, US, and New Delhi, India, have densities of 7,101 and 5,855 inhabitants per squared kilometre, respectively.

Because of the multiple factors influencing the diffusion rate of the virus, the geographical spread is heterogeneous, with some neighbourhoods having a higher incidence than others. From a public health perspective, predicting neighbourhoods at high risk of incidence for severe COVID-19 infections is particularly useful to organize targeted actions and to reduce hospital overcrowding and patient burden in such areas. While clinical factors can easily be retrieved from electronic health records, socio-economic determinants are not always available but can be gathered from census data available through patient geolocalization. In most countries, census data enable the assessment of a large variety of socio-economic indicators at fine geographical levels, and many ecological studies have already been performed on COVID-19 using these data (2,3). For instance, in patients from the UK biobank, a striking gradient in COVID-19 hospitalization rates according to the Townsend Deprivation Index - a composite measure of socio-economic deprivation - and household income was observed (4). In the US, wide inequities have been shown (5) in COVID-19 positivity and incidence, with less advantaged neighbourhoods having a higher incidence in Chicago, NYC and Philadelphia. In the megacity of Chennai (India) (6), spatial risks of COVID-19 were predominantly concentrated in wards with higher levels of area deprivation, which were mostly located in the northeast parts of Chennai. The same holds for England and Wales (7). Other indicators among the many analysed have been shown to be associated with COVID-19 incidence, such as ethnicity (8) and population density (9).

Previous studies only analysed a few indicators together and showed that most of them were associated with COVID-19 incidence. A socio-economic environment-wide analysis considering a very large set of socio-economic indicators would enable a global picture of neighbourhoods at risk. This data-mining strategy has become common since the first genome-wide association studies (10) and has been adapted to many kinds of data, such as phenotypes (11) and medications (12). This type of study would enable the pinpointing of specific environmental characteristics strongly associated with COVID-19 risk with no *a priori* hypothesis regarding the type of associated indicator. Using highly associated characteristics, a predictive model could be inferred to precisely highlight neighbourhoods at risk for a greater incidence of severe COVID-19.

In this paper, our objective is to perform a socio-economic environment-wide study and to analyse to what extent a large set of census indicators can predict the incidence of hospitalization for COVID-19 in the Paris area.

## Materials and Methods

Our analysis follows recommendations provided by the REporting of studies Conducted using Observational Routinely-collected health Data (RECORD) Statement. Completed checklist is available as supplementary material (Table S2). The socio-economic wide association study method presented here aims at testing the association between a large set of socio-economic indicators from French census data and COVID-19 hospitalization risk.

### Data extraction

The AP-HP is the largest hospital entity in Europe, with 39 hospitals (22,474 beds) mainly located in the Greater Paris area, with 1.5 M hospitalizations per year (10% of all hospitalizations in France). Because of the centralized healthcare organization during the COVID epidemic, COVID-infected patients were mostly hospitalized in AP-HP hospitals, and private clinics were not habilitated to take charge of COVID patients.

Since 2014, the AP-HP has deployed an analytics platform based on a clinical data repository (CDR), aggregating day-to-day clinical data from 8.8 million patients captured by clinical databases. An “EDS-COVID” database stemmed from this initiative. The AP-HP COVID database retrieved electronic health records from all AP-HP facilities and aggregated them into a clinical data warehouse following the OMOP common data model (13). The clinical data warehouse allows a large set of data to be retrieved in real time to deeply characterize hospitalized patients, including their residential address. Using the IRIS Paris division and adopting one-week periods, we aggregated the data for the new patients who tested positive for COVID-19 in one of the AP-HP facilities and lived in Paris.

IRIS, as defined by the National Institute for Statistics and Economic Studies (INSEE), is the smallest geographical division in France, with 2000 inhabitants, on average (towns with more than 5000 inhabitants are divided into several IRIS, while smaller towns form one IRIS each). There are two types of IRIS: business and residential. The homogeneity of each unit is based mainly on habitat type (residential area, public housing, etc.) Therefore, each French residential IRIS has relatively homogeneous social characteristics.

New patients who tested positive by polymerase chain reaction (PCR) as being infected by SARS-CoV-2, hospitalized in one of the AP-HP hospitals and living in Paris were aggregated at the IRIS Paris division level to constitute the dataset for this study.

For each of the INSEE residential units, INSEE provides several socio-economic ecological open datafiles stemming from census data (14). For each IRIS unit, we retrieved all information available as open data at this IRIS scale: 71 indicators on population demographics and distribution, 51 on housing, 86 on professional activity, 55 on family types, 19 on education and 21 on income, for a total of 303 indicators.

### Methods

For each Paris IRIS residential unit, we estimated the proportion of patients hospitalized at AP-HP from the 1^st^ of February until the 4^th^ of June. High- and low-risk IRIS units were defined using the first decile of this proportion. We then assessed the association of a given socio-economic indicator above median value with having a high COVID-19 risk for each of the 303 socio-economic indicators using a chi-square test. This enables an equivalent power when testing the association with each indicator. This is an important point of the methodology of this study, as it enables us to directly compare the magnitude of odds ratios for each indicator. P-values for the different performed tests were represented through a Manhattan plot using the indicator type as the variable group.

We randomly divided the IRIS residential unit into a training (66%) and a test (33%) dataset.

In the training set, we selected for the prediction model all indicators reaching significance at the 0.1% level and having a correlation between each other lower than 80%. To achieve this selection, we ranked indicators reaching significance at 0.1% from the lower to the higher p-values, and we withdrew each indicator presenting a correlation larger than 80% with one of the other indicators. We then performed a multivariate regularized logistic regression using lasso cross-validated penalty, with the selected indicators as the explaining variable and COVID-19 risk as the variable to explain.

We estimated the prediction performance of the fitted model on the training set by estimating the area under the curve (AUC). We simplified the full model when relevant: for each included variable in this model, we also estimated the corresponding AUC and compared it with the AUC of the full model using the Delong method. For the final model, we then estimated the threshold maximizing the Youden index and the corresponding sensitivity and specificity.

We applied the final model to the test set and estimated the AUC and the sensitivity and specificity corresponding to the above estimated Youden index.

To take into account possible spatial autocorrelation, we tested spatial autocorrelation in the final model residuals using the Moran’s I test.

All analyses were performed using R software with the ggplot, pROC and DHARMa packages.

## Results

As of the 3rd of June 2020 (the end of the first wave of the epidemic), 22,132 patients had tested as positive in one of the AP-HP facilities; among them, 18,634 had their residential address geolocalized in the Ile-de-France region. Among these 18,634 patients, 7409 had a residential address in Paris, of whom 6563 patients had an address in one of the 855 Paris residential areas, i.e., with available census indicators (figure 1). Among these 6563 patients, 3400 patients were hospitalized in one the the 855 residential areas, with a median number of hospitalized cases per residential areas of 3 cases (IQR95%: [1; 5]). The first decile of COVID-19 hospitalization rate among Paris residential areas was equal to 3 per 1000 inhabitants. We dichotomized the Paris residential areas below and above this threshold.

**Figure 1:**
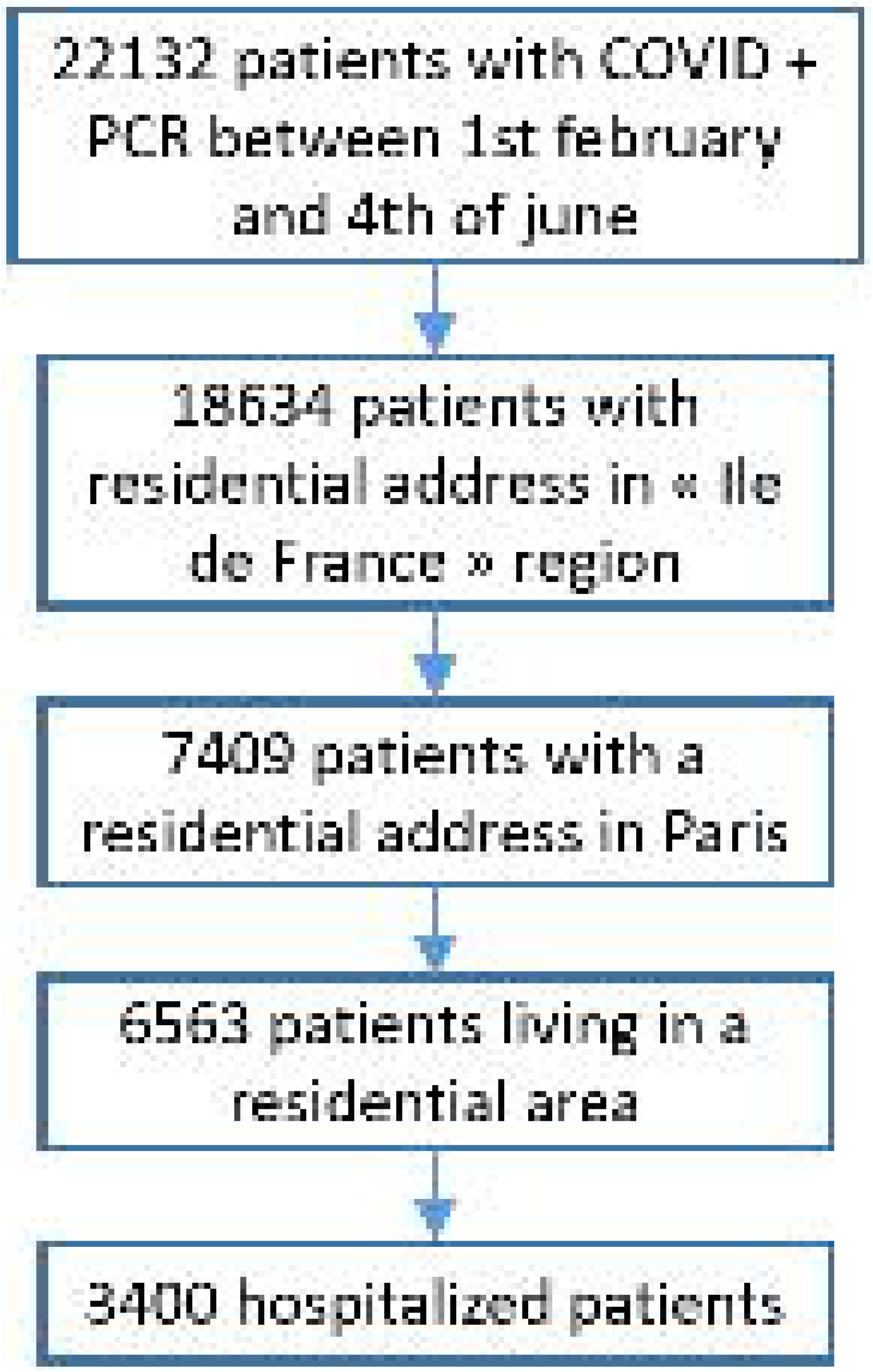
Population flow chart.

Many indicators were strongly associated with hospitalization rate (figure 2, table S3), the most strongly associated indicators being related to income and occupational status. Regarding income, the most significantly associated indicator was income corresponding to the 3^rd^ decile of the population (OR= 0.11, CI95% [0.05; 0.20]). Regarding population indicators, the most significantly associated indicators concerned population activities, with the rate of workers in blue-collar unskilled occupations being strongly associated with high-risk areas (OR=4.38, CI95% [2.67; 7.51]) and the highly qualified worker rate being strongly associated with low-risk areas (OR=0.19, CI95% [0.10; 0.31]). Population demographics were much less associated, with the highest association reached by the rate of women aged between 45 and 59 years (OR=2.17, CI95% [1.40;3.44]). Of note, population density was not associated, and the immigrant rate was strongly associated (OR=3.20, CI95% [2.01; 5.25]). Regarding population occupational status, the rate of active workers in unskilled occupations was strongly associated with high-risk areas (OR=5.04, CI95% [3.03; 8.85]), and the active population rate in the 15-24 year age group was strongly associated with low-risk areas (OR=0.21, CI95% [0.12; 0.35]). Another indicator that was strongly associated with high-risk areas was the unemployment rate (OR=4.69, CI95% [2.84; 8.14]). Regarding housing, the most strongly associated indicators were the primary resident rate (OR=5.87, CI95% [3.46;10.61]) and public low-rent housing rate (OR=4.69, CI95% [2.84; 8.14]). Regarding education, the most associated indicator was the rate of women over 15 years old with no diploma (OR= 5.04, CI95% [3.03;8.85]), and the rate of women with high education was associated with low-risk areas (OR=0.19, CI95% [0.10; 0.31]). Regarding households, the two most associated indicators were the rate of household heads with an unskilled occupation (OR=4.38, CI95% [2.67; 7.51]) and the rate of single-parent families (OR=3.84, CI95% [2.37;6.46]).Figure 3 represents the gradient of the 3^rd^ decile income of the population area and the hospitalization rate in Paris. These maps taken together confirm the strong association between these two variables.

**Figure 2:**
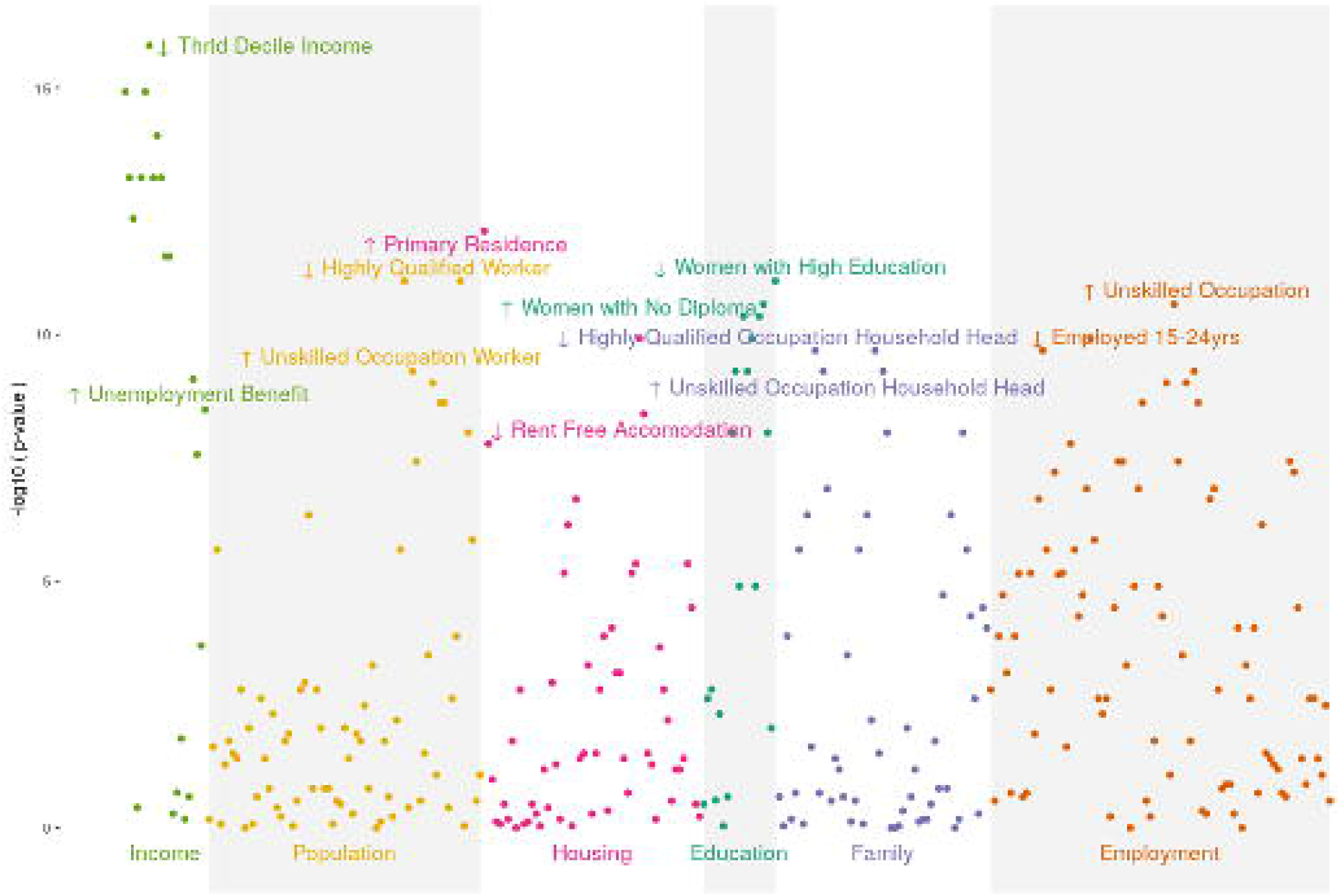
Manhattan plot representing the p-value of the 303 association tests between the COVID-19 hospitalization rate and each socio-economic factor.

**Figure 3:**
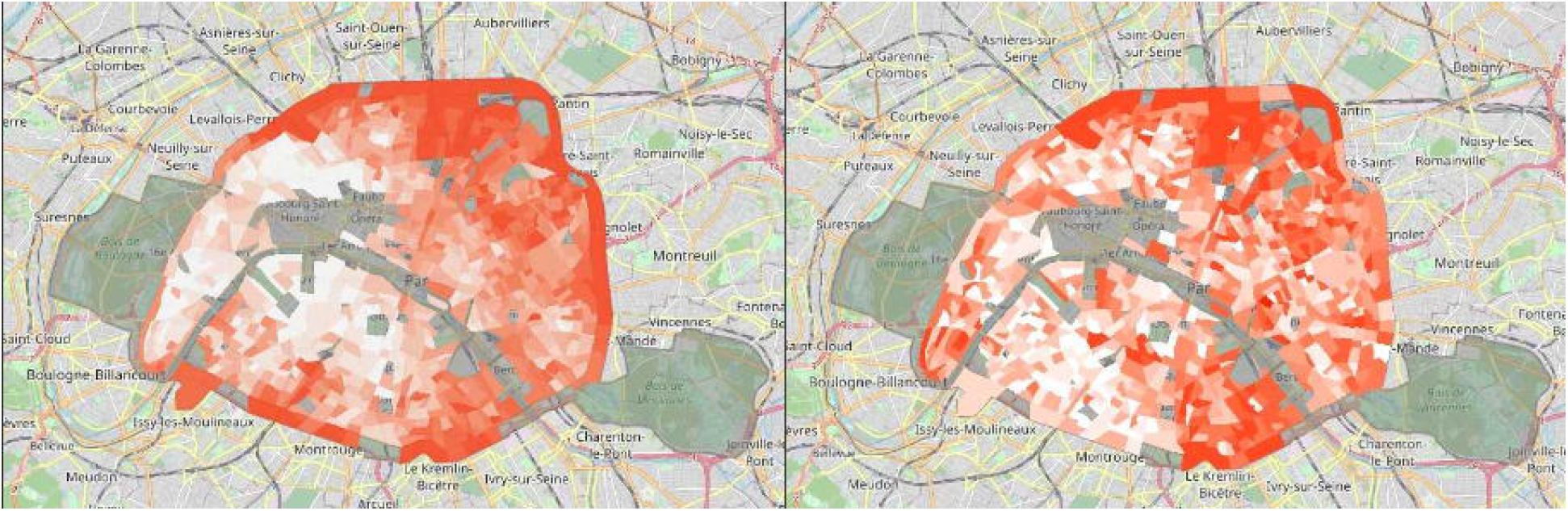
Gradient of income third decile (left) and COVID-19 hospitalized cases >3 per 1000 inhabitants (right) in Paris residential areas. Non-residential areas are not covered by a red-colour gradient.

To build a predictive model, Paris residential areas were split into a training set of 570 IRIS residential areas and a test set of 285 IRIS residential areas. Of the 303 available indicators, 104 were significantly associated with the hospitalization rate at the 0.1% level, resulting in 29 independent associated features after removing strongly correlated variables (figure 4). The three independently associated indicators selected by the regularized multivariate regression are the income corresponding to the 3^rd^ decile of the population area (adjusted OR= 0.22, CI95% [0.09; 0.52]), the rate of highly qualified working women (adjusted OR=0.33, CI95% [0.16; 0.70]), and the rate of women over 15 years old with no diploma (adjusted OR=1.90, CI95% [0.82; 4.43]), leading to an estimated AUC of 0.77 (CI 95%: 0.75-0.86). We then considered each of these indicators as continuous. This led to an AUC of 0.80 (CI 95%: 0.73-0.83). When using the Delong test to compare the multivariate model with these 3 indicators and each indicator taken separately, there was no significant difference between the full model and the models considering each indicator taken separately. We therefore end up with a final model containing only the income corresponding to the 3^rd^ decile of the population (AUC=0.78, CI95%: 0.72-0.85). The final model residuals did not show spatial autocorrelation (Moran’s I test, p=0.66). The corresponding sensitivities and specificities for the maximal value of the Youden index were 84% (CI95%: 0.73-0.92) and 62% (CI95%: 0.58-0.66) for a cut-off value of 20,300€, with 43% (CI95%: 0.39-0.47) of residential IRIS areas below this threshold (table 1). In the test set, the AUC was similar (AUC=0.79 (CI95%: 0.71-0.87), and the 50% most deprived areas concentrate 90% of the areas with high COVID-19 prevalence.

**Table 1:** Performance obtained for the final model for the training and the test set using the third decile of income, 20,300€, as the threshold

|  | *Training set* | *Test set* |
| --- | --- | --- |
| *Number of Paris residential areas* | 570 | 285 |
| *True positives* | 54 | 28 |
| *False positives* | 191 | 114 |
| *True negatives* | 10 | 3 |
| *False negatives* | 315 | 140 |
| *Prevalence* | 0.43 (0.39, 0.47) | 0.50 (0.44, 0.56) |
| *Sensitivity* | 0.84 (0.73, 0.92) | 0.90 (0.74, 0.98) |
| *Specificity* | 0.62 (0.58, 0.66) | 0.55 (0.49, 0.61) |
| *Positive predictive value* | 0.22 (0.17, 0.28) | 0.20 (0.14, 0.27) |
| *Negative predictive value* | 0.97 (0.94, 0.99) | 0.98 (0.94, 1.00) |
| *Positive likelihood ratio* | 2.24 (1.92, 2.61) | 2.01 (1.68, 2.41) |
| *Negative likelihood ratio* | 0.25 (0.14, 0.45) | 0.18 (0.06, 0.52) |

**Figure 4:**
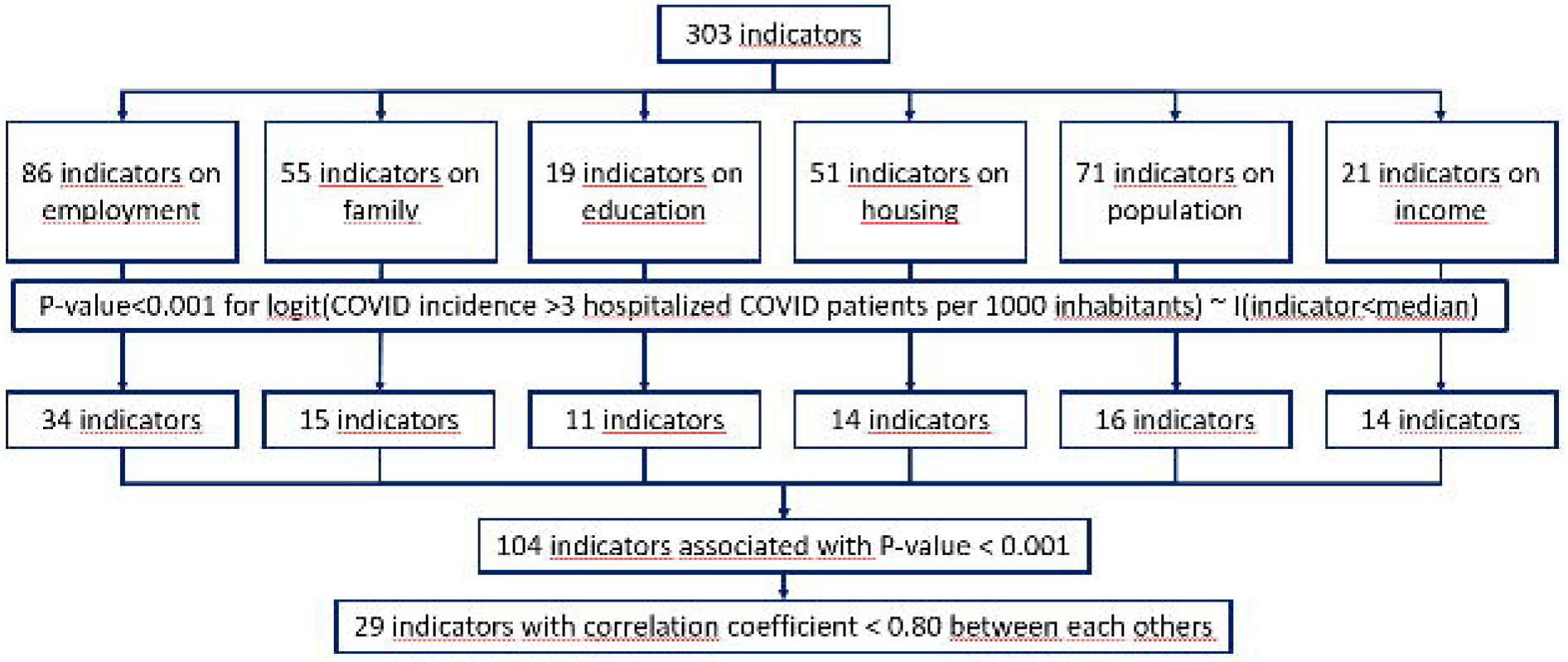
Flowchart for indicators’ selection in the predictive model using the training sample.

## Discussion

Using a high-dimensional set of socio-economic indicators, we show that many socio-economic indicators are strongly associated with an area of high incidence of hospitalized patients for COVID-19 in Paris, with an odds ratio of magnitude of approximately 5 for indicators measuring deprivation. As a consequence, several of these indicators, such as income, proportion of the population with no diploma, and the rate of workers in unskilled occupations, are by themselves highly predictive of hospitalized case clusters. Overall, the 45% most deprived areas comprised 86% of the areas with a high incidence of COVID-19 hospitalized cases.

This type of study, collecting passively data from hospitalized cases, therefore not necessitating any effort for health caregivers, is of particular interest in such a period of hospital overcrowding.Indeed, it can provide real-time information on populations having high COVID-19 incidence. This type of study, with nearly no cost, has to be faced to seroprevalence studies deemed to be biased due to non-respondents, involving huge research efforts and not useable for real-time surveillance.

This study highlights the high pertinence of geographic breakdown at the small IRIS level that is finely described using census data. This can both preserve patients’ privacy and finely depict patients’ environment. Indeed, geolocalization based on the IRIS area level renders the data anonymous while being pertinent to describing patients’ socio-economic environment, as attested by the strong value of the predictive model built using these data.

This study is based on declared residential addresses, which can be different from a patient’s living residence at the time of hospitalization. Interestingly, 14% of hospitalized patients have their residential address outside the Ile-de-France region and are therefore located far from Paris.Although some transfers from remote hospitals occurred during the study period, we cannot exclude the possibility that some patients were actually living in Paris at the time of their hospitalization. The same holds for the considered indicators, which were based on census data that might differ during the study period because a large exodus from Paris was observed when lockdown started: during lockdown, in addition to the departure of foreign visitors and overseas departments, Paris saw its current population decrease by 450,000 people (i.e., 20%). Half of this population decline was due to non-residents of the capital who were able to return home, and the other half was due to Parisians leaving their city (15).

Only public hospitals were considered for this study. This is unlikely to have introduced a bias because private facilities were not allowed to take charge of patients affected by COVID-19, with the exception of non-profit private facilities, in which patients were hospitalized according to the same financial conditions as public hospitals. Moreover, non-profit private hospitals are distributed evenly across Paris like public hospitals are.

A significant portion of hospitalized patients came from medical institutions, such as medico-welfare and long-term care establishments. These patients were not considered in our study, as the vast majority of these establishments are located in non-residential areas. Considering such patients in our study would have introduced a bias as the transmission mechanism underlying these cases is different from the general population.

Notably, we did not include air pollution data because they were not available at the same scale. Some studies have already shown that air pollution might increase the COVID-19 mortality rate (16,17), but these studies did not consider socio-economic determinants, while low-income areas are usually located in more polluted areas (18). In Paris, low-income areas are concentrated near the Parisian ring road, which is a highly polluted area. Therefore, it would have been interesting to have these data considered in our model. Moreover, from the map, it can visually be deduced that income is a more important determinant than ring road proximity.

The predictive value of low-income neighbourhoods on epidemic clusters is high and can be explained both by factors favouring infection, such as occupational status, and by factors favouring severe infection in infected patients, such as obesity and hypertension. The subgroup pinpointed as overrepresented in the high-risk area, i.e., the population with unskilled occupations living in deprived areas, is not considered to be at high risk of severe COVID-19 because these people are still active, i.e., below 65 years, and age is the major risk factor for severe COVID-19 infection. However, they might have a high rate of infection because of their professional status (residence employee, housekeeper, delivery driver, etc.) involving a large number of professional contacts with the more vulnerable population. This strong predictive value of income level for areas at high risk has to be compared with the low level of inequalities in France compared to the level worldwide (19). The French health care system is ranked among the better regarding waiting times, results, and generosity (20), which renders these results unexpected. Indeed, many studies have performed ecological analysis on COVID-19 mortality and hospitalization rates but have not found such a strong association in other countries, including the US and the United Kingdom, but their study design was slightly different, with an odds ratio of approximately 3 for poverty in Chicago (21) and of approximately 2 in England (1), for instance, while odds ratios were as high as 5 in our study.

We did not find any association with population density. The population density in residential areas is high (37,000 inhabitants per km2 in residential areas), but the interquartile range is also high (from 28,000 to 46,000 inhabitants per km2 in residential areas). Therefore, this lack of association is not explained by a homogeneous population density. In Paris, population density is not strongly correlated with population wealth, as opposed to the case in most other large cities, where parts with high densities correspond to low-income neighbourhoods (21,22). This might explain why population density was associated with high COVID-19 incidence in other areas but not in Paris.

Moreover, most previous studies focus on COVID-19 incidence, sometimes in relationship with testing incidence (5). COVID-19 incidence strongly depends on testing policy, which is associated with socio-economic determinants of the population. Therefore, analysing the association between COVID-19 incidence and socio-economic determinants is deemed to be biased. This is why we focused on COVID-19 hospitalized cases, which better reflect epidemic spread because they are less related to testing policy. This was particularly relevant in this study, as a low-income population might have difficulties accessing testing facilities (23) because of the testing cost and long working hours of this population.

This study only includes data from the first wave of the epidemic. Different mechanisms could be at stake in the future. In Germany, it was shown that different mechanisms were responsible for epidemic spread at different stages of the epidemic (24). The period analysed in this study corresponds mostly to lockdown, which has a strong impact on population behaviour in France and therefore might have led to very specific mechanisms for epidemic spread. Nevertheless, our study highlighted that the proposed methodology in this paper, a socio-economic wide association study in hospitalized patients, can provide indirect information on populations having high COVID-19 incidence. It is therefore of utmost importance to organize a real-time follow-up of these socio-economic indicators for newly hospitalized patients using such an approach.

## Data Availability

Aggregated data at IRIS level and programming codes are available on request from the corresponding author after approval by the institutional review board (authorization number IRB 00011591) from the Scientific and Ethical Committee from the AP-HP (Assistance Publique Hopitaux de Paris) for research purposes in line with French health data regulation.

## Acknowledgements

We would like to acknowledge the authors thank the EDS APHP Covid consortium integrating the APHP Health Data Warehouse team as well as all the APHP staff and volunteers who contributed to the implementation of the EDS-COVID database and operating solutions for this database (list in the Supplementary Table S1).

## Author approval

All authors have seen and approved the manuscript.

## Ethics approval

This study was approved by the institutional review board (authorization number IRB 00011591) from the Scientific and Ethical Committee from the AP-HP (Assistance Publique – Hôpitaux de Paris).

## Consent to participate

All subjects included in this study were informed about the reuse of their data for research, and subjects who objected to the reuse of their data were excluded from this study, in accordance with French legislation.

## Availability of data and material (data transparency)

Aggregated data at IRIS level and programming codes are available on request from the corresponding author after approval by the institutional review board (authorization number IRB 00011591) from the Scientific and Ethical Committee from the AP-HP (Assistance Publique – Hôpitaux de Paris) for research purposes in line with French health data regulation.

## Code availability (software application or custom code)

Custom codes are available on request from the corresponding author.

## Contributions

ASJ designed the study. HC and BR performed the geolocalization of patients’ residential address and the data aggregation. ASJ, HC and BR had full access to aggregated data used for this study and take responsibility for the integrity of the data. ASJ did the analyses and takes responsibility for the accuracy of the data analysis. ASJ drafted the paper with the help of BR, AB, SK. Data were collected from all Assistance Publique - Hôpitaux de Paris. All authors critically revised the manuscript for important intellectual content and gave final approval for the version to be published.

