## Supplemental materials for "COVID-19, a social disease in Paris: a socio-economic wide association study on hospitalized patients highlights low-income neighbourhood as a key determinant of severe COVID-19 incidence during the first wave of the epidemic"

### Table S1. List of collaborators

| Nom | Prénom | Affiliation | Contribution |
|---|---|---|---|
| Ancel | Pierre-Yves | APHP Paris University Center | Local CDW coordinator |
| Bauchet | Alain | AP-HP Saclay University | |
| Beeker | Nathanael | AP-HP Paris University Center | Data scientist |
| Benoit | Vincent | WIND Department APHP Greater Paris University Hospital | Data engineer |
| Bernaux | Mélodie | Strategy and transformation department, APHP Greater Paris University Hospital | Medical coordination of data analysis |
| Bellamine | Ali | WIND Department APHP Greater Paris University Hospital | Data engineer, data scientist |
| Bey | Romain | WIND Department APHP Greater Paris University Hospital | Data engineer, data scientist, regulatory assessment |
| Bourmaud | Aurélie | APHP Paris University North | Local CDW coordinator |
| Bréant | Stéphane | WIND Department APHP Greater Paris University Hospital | Coordination of clinical research informatics |
| Burgun | Anita | Department of Biomedical Informatics, HEGP, APHP Greater Paris University Hospital | Medical & scientific coordination |
| Carrat | Fabrice | APHP Sorbonne University | |
| Caucheteux | Charlotte | Université Paris-Saclay, Inria, CEA | Data integration and analysis |
| Champ | Julien | INRIA Sophia-Antipolis – ZENITH team, LIRMM, Montpellier, France | Data integration and analysis |
| Cormont | Sylvie | WIND Department APHP Greater Paris University Hospital | Data standardisation |
| Daniel | Christel | WIND Department APHP Greater Paris University Hospital UMRS1142 INSERM | Medical director of data standardisation and clinical research infor[illegible] |
| Dubiel | Julien | WIND Department APHP Greater Paris University Hospital | Data engineer |
| Ducloas | Catherine | APHP Paris Seine Saint Denis Universitary Hospital | Local CDW coordinator |
| Esteve | Loic | SED/SIERRA, Inria Centre de Paris | Data engineer, data scientist |
| Frank | Marie | APHP Saclay University | Local CDW coordinator |
| Garcelon | Nicolas | Imagine Institute | Data engineer, data scientist |
| Gramfort | Alexandre | Université Paris-Saclay, Inria, CEA | Data engineer, data scientist |
| Griffon | Nicolas | WIND Department APHP Greater Paris University Hospital, UMRS1142 INSERM | Data standardisation |
| Grisel | Olivier | Université Paris-Saclay, Inria, CEA | Data engineer, data scientist |
| Guilbaud | Martin | WIND Department APHP Greater Paris University Hospital | Data engineer |
| Hassen-Khodja | Claire | Direction of the Clinical Research and Innovation, AP-HP | Medical coordination of data-driven research |
| Hemery | François | APHP Henri Mondor University Hospital | Local CDW coordinator |
| Hilka | Martin | WIND Department APHP Greater Paris University Hospital | Director of Big data platform |
| Jannot | Anne Sophie | Department of Biomedical Informatics, HEGP, APHP Greater Paris University Hospital | Biostatistician, local CDW coordonator |

| | | | |
|---|---|---|---|
| Lambert | Jerome | APHP Paris University North | Local CDW coordinator |
| Layese | Richard | APHP Henri Mondor University Hospital | |
| Leblanc | Judith | Clincial Research Unit, Saint Antoine Hospital, APHP Greater Paris University Hospital | Data scientist |
| Lebouter | Léo | WIND Department APHP Greater Paris University Hospital | Data engineer |
| Lemaitre | Guillaume | Université Paris-Saclay, Inria, CEA | Data engineer, data scientist |
| Leprovost | Damien | Clevy.io | Data engineer, data scientist |
| Lerner | Ivan | Department of Biomedical Informatics, HEGP, APHP Greater Paris University Hospital | Data engineer, data scientist |
| Levi Sallah | Kankoe | APHP Paris University North | |
| Maire | Aurélien | WIND Department APHP Greater Paris University Hospital | Data engineer |
| Mamzer | Marie-France | President of the AP-HP IRB | President of the AP-HP IRB |
| Martel | Patricia | APHP Saclay University | Data scientist |
| Mensch | Arthur | ENS, PSL University | Data engineer, data scientist |
| Moreau | Thomas | Université Paris-Saclay, Inria, CEA | Data engineer, data scientist |
| Neuraz | Antoine | Department of Biomedical Informatics, HEGP, APHP Greater Paris University Hospital | Data engineer, data scientist |
| Orlova | Nina | WIND Department APHP Greater Paris University Hospital | Data engineer |
| Paris | Nicolas | WIND Department APHP Greater Paris University Hospital | Data engineer, data scientist |
| Rance | Bastien | Department of Biomedical Informatics, HEGP, APHP Greater Paris University Hospital | Data engineer, data scientist |
| Ravera | Hélène | WIND Department APHP Greater Paris University Hospital | Data engineer |
| Rozes | Antoine | APHP Sorbonne University | |
| Salamanca | Elisa | WIND Department APHP Greater Paris University Hospital | Director of the Data & Innovation department |
| Sandrin | Arnaud | WIND Department APHP Greater Paris University Hospital | Director of the National Rare Diseases Database |
| Serre | Patricia | WIND Department APHP Greater Paris University Hospital | Data engineer, data standardisation |
| Tannier | Xavier | Sorbonne University | Data engineer, data scientist |
| Treluyer | Jean-Marc | APHP Paris University Center | Local CDW coordinator |
| Van Gysel | Damien | APHP Paris University North | Local CDW coordinator |
| Varoquaux | Gael | Université Paris-Saclay, Inria, CEA, Montréal Neurological Institute, McGill University | Data engineer, data scientist |
| Vie | Jill Jen | SequeL, Inria Lille | Data engineer, data scientist |
| Wack | Maxime | Department of Biomedical Informatics, HEGP, APHP Greater Paris University Hospital | Data engineer, data scientist |
| Wajsburt | Perceval | Sorbonne University | Data engineer, data scientist |

| | | | |
|---|---|---|---|
| Wassermann | Demian | Université Paris-Saclay, Inria, CEA | Data engineer, data scientist |
| Zapletal | Eric | Department of Biomedical Informatics, HEGP, APHP Greater Paris University Hospital | Data engineer |

**Collégiales of AP-HP:** anesthésie-réanimation, médecine intensive réanimation, infectiologie, virologie, nutrition

**Table S2: The RECORD statement – checklist of items, extended from the STROBE statement, that should be reported in observational studies using routinely collected health data.**

| | Item No. | STROBE items | Location in manuscript where items are reported | RECORD items | Location in manuscript where items are reported |
|---|---|---|---|---|---|
| **Title and abstract** | | | | | |
| | 1 | (a) Indicate the study's design with a commonly used term in the title or the abstract (b) Provide in the abstract an informative and balanced summary of what was done and what was found | | RECORD 1.1: The type of data used should be specified in the title or abstract. When possible, the name of the databases used should be included.<br><br>RECORD 1.2: If applicable, the geographic region and timeframe within which the study took place should be reported in the title or abstract.<br><br>RECORD 1.3: If linkage between databases was conducted for the study, this should be clearly stated in the title or abstract. | Specified in abstract<br><br>Specified both in title and in abstract<br><br>No linkage |
| **Introduction** | | | | | |
| Background rationale | 2 | Explain the scientific background and rationale for the investigation being reported | | | Specified |
| Objectives | 3 | State specific objectives, including any prespecified hypotheses | | | Specified |
| **Methods** | | | | | |
| Study Design | 4 | Present key elements of study | | | Specified at |

| | | | | | |
|---|---|---|---|---|---|
| | | design early in the paper | | | the beginning of materials and methods section |
| Setting | 5 | Describe the setting, locations, and relevant dates, including periods of recruitment, exposure, follow-up, and data collection | | | Describe in data extraction section |
| Participants | 6 | *(a) Cohort study* - Give the eligibility criteria, and the sources and methods of selection of participants. Describe methods of follow-up<br>*Case-control study* - Give the eligibility criteria, and the sources and methods of case ascertainment and control selection. Give the rationale for the choice of cases and controls<br>*Cross-sectional study* - Give the eligibility criteria, and the sources and methods of selection of participants<br><br>*(b) Cohort study* - For matched studies, give matching criteria and number of exposed and unexposed | | RECORD 6.1: The methods of study population selection (such as codes or algorithms used to identify subjects) should be listed in detail. If this is not possible, an explanation should be provided.<br><br>RECORD 6.2: Any validation studies of the codes or algorithms used to select the population should be referenced. If validation was conducted for this study and not published elsewhere, detailed methods and results should be provided.<br><br>RECORD 6.3: If the study involved linkage of databases, consider use of a flow diagram or other graphical display to demonstrate the data linkage process, including the number of individuals with linked data at each stage. | Describe in data extraction section<br><br>Not relevant<br><br>Figure 1 |

| | | | | | |
|---|---|---|---|---|---|
| | | *Case-control study* - For matched studies, give matching criteria and the number of controls per case | | | |
| Variables | 7 | Clearly define all outcomes, exposures, predictors, potential confounders, and effect modifiers. Give diagnostic criteria, if applicable. | | RECORD 7.1: A complete list of codes and algorithms used to classify exposures, outcomes, confounders, and effect modifiers should be provided. If these cannot be reported, an explanation should be provided. | Given in supplementary table S3 |
| Data sources/ measurement | 8 | For each variable of interest, give sources of data and details of methods of assessment (measurement). Describe comparability of assessment methods if there is more than one group | | | Given in data extraction section<br><br>Given in stantistical analysis section |
| Bias | 9 | Describe any efforts to address potential sources of bias | | | Describe in data extraction section |
| Study size | 10 | Explain how the study size was arrived at | | | Describe in figure 1 |
| Quantitative variables | 11 | Explain how quantitative variables were handled in the analyses. If applicable, describe which groupings were chosen, and why | | | Describe in statistical analysis section |
| Statistical methods | 12 | (a) Describe all statistical methods, including those used to control for confounding | | | Describe in statistical analysis section |

| | | | | | |
|---|---|---|---|---|---|
| | | (b) Describe any methods used to examine subgroups and interactions<br>(c) Explain how missing data were addressed<br>(d) *Cohort study* - If applicable, explain how loss to follow-up was addressed<br>*Case-control study* - If applicable, explain how matching of cases and controls was addressed<br>*Cross-sectional study* - If applicable, describe analytical methods taking account of sampling strategy<br>(e) Describe any sensitivity analyses | | | No interaction test<br><br>No missing data<br><br>No follow-up |
| Data access and cleaning methods | | .. | | RECORD 12.1: Authors should describe the extent to which the investigators had access to the database population used to create the study population.<br><br>RECORD 12.2: Authors should provide information on the data cleaning methods used in the study. | Describe in the contribution section and data extraction section |
| Linkage | | .. | | RECORD 12.3: State whether the study included person-level, institutional-level, or other data linkage across two or more databases. The methods of linkage and methods of linkage quality | Describe in data extraction section |

| | | | | | |
|---|---|---|---|---|---|
| | | | | evaluation should be provided. | |
| **Results** | | | | | |
| Participants | 13 | (a) Report the numbers of individuals at each stage of the study (*e.g.*, numbers potentially eligible, examined for eligibility, confirmed eligible, included in the study, completing follow-up, and analysed)<br>(b) Give reasons for non-participation at each stage.<br>(c) Consider use of a flow diagram | | RECORD 13.1: Describe in detail the selection of the persons included in the study (*i.e.*, study population selection) including filtering based on data quality, data availability and linkage. The selection of included persons can be described in the text and/or by means of the study flow diagram. | A flow diagram is provided (figure 1) |
| Descriptive data | 14 | (a) Give characteristics of study participants (*e.g.*, demographic, clinical, social) and information on exposures and potential confounders<br>(b) Indicate the number of participants with missing data for each variable of interest<br>(c) *Cohort study* - summarise follow-up time (*e.g.*, average and total amount) | | | No analysis at patient-level. Indicators at IRIS-level are described in table S3<br><br>No follow-up. |
| Outcome data | 15 | *Cohort study* - Report numbers of outcome events or summary measures over time<br>*Case-control study* - Report numbers in each exposure category, or | | | Number of hospitalized cases per IRIS is described |

| | | | | | |
|---|---|---|---|---|---|
| | | summary measures of exposure *Cross-sectional study* - Report numbers of outcome events or summary measures | | | |
| Main results | 16 | (a) Give unadjusted estimates and, if applicable, confounder-adjusted estimates and their precision (e.g., 95% confidence interval). Make clear which confounders were adjusted for and why they were included<br>(b) Report category boundaries when continuous variables were categorized<br>(c) If relevant, consider translating estimates of relative risk into absolute risk for a meaningful time period | | | Reported in Table S3 |
| Other analyses | 17 | Report other analyses done—e.g., analyses of subgroups and interactions, and sensitivity analyses | | | No other analysis |
| **Discussion** | | | | | |
| Key results | 18 | Summarise key results with reference to study objectives | | | Reported in the beginning of the discussion section |
| Limitations | 19 | Discuss limitations of the study, taking into account sources of potential | | RECORD 19.1: Discuss the implications of using data that were not | Included in discussion section |

| | | | | | |
|---|---|---|---|---|---|
| | | bias or imprecision. Discuss both direction and magnitude of any potential bias | | created or collected to answer the specific research question(s). Include discussion of misclassification bias, unmeasured confounding, missing data, and changing eligibility over time, as they pertain to the study being reported. | |
| Interpretation | 20 | Give a cautious overall interpretation of results considering objectives, limitations, multiplicity of analyses, results from similar studies, and other relevant evidence | | | Included in discussion section |
| Generalisability | 21 | Discuss the generalisability (external validity) of the study results | | | Included at the end of the discussion section |
| **Other Information** | | | | | |
| Funding | 22 | Give the source of funding and the role of the funders for the present study and, if applicable, for the original study on which the present article is based | | | Included |
| Accessibility of protocol, raw data, and programming code | | .. | | RECORD 22.1: Authors should provide information on how to access any supplemental information such as the study protocol, raw data, or programming code. | Included |

*Checklist is protected under Creative Commons Attribution (CC BY) license.

Table S3: Unit, median value and interquartile range (IQR), odds-ratios (OR) and their confidence intervals at 95% (CI95%) corresponding to each indicator. Because by construction, sample sizes are the same for all indicators, they are not given.

Income indicators

| *Indicator* | *Units* | *Median [IQR]* | *OR* | *CI95%* |
|---|---|---|---|---|
| *First quartile* | k€ | 18 [13;23] | 0.12 | [0.06;0.22] |
| *Median value* | k€ | 31 [25;39] | 0.15 | [0.08;0.26] |
| *Third quartile* | k€ | 48 [38;60] | 0.16 | [0.09;0.28] |
| *Interquartile range relative to the median value* | % | 103 [92;114] | 1.21 | [0.79;1.86] |
| *First decile* | k€ | 8 [6;11] | 0.15 | [0.08;0.26] |
| *Second decile* | k€ | 15 [11;19] | 0.12 | [0.06;0.22] |
| *Third decile* | k€ | 21 [15;26] | 0.11 | [0.05;0.2] |
| *Fourth decile* | k€ | 26 [20;32] | 0.15 | [0.08;0.26] |
| *Sixth decile* | k€ | 37 [29;46] | 0.13 | [0.07;0.24] |
| *Seventh decile* | k€ | 44 [35;55] | 0.15 | [0.08;0.26] |
| *Eighth decile* | k€ | 53 [42;68] | 0.17 | [0.1;0.3] |
| *Ninth decile* | k€ | 70 [54;92] | 0.17 | [0.1;0.3] |
| *Interdecile ratio* | - | 9 [8;10] | 0.86 | [0.56;1.32] |
| *Ratio 80%/20% by consumption unit* | - | 12 [10;14] | 0.75 | [0.48;1.15] |
| *Gini index by consumption unit* | % | 42 [39;46] | 0.57 | [0.35;0.89] |
| *In-work income rate* | % | 75 [70;80] | 1.1 | [0.72;1.69] |
| *Salaried income rate* | % | 66 [61;71] | 1.31 | [0.85;2.02] |
| *Unemployment benefit rate* | % | 3 [2;4] | 4.3 | [2.64;7.31] |
| *Non-salaried income rate* | % | 6 [4;8] | 0.26 | [0.16;0.43] |
| *Retirement and pension benefit rate* | % | 17 [14;21] | 2.31 | [1.48;3.68] |
| *Other income rate* | % | 6 [4;9] | 0.24 | [0.14;0.39] |

Population indicators

| *Indicators* | *Units* | *Median [IQR]* | *OR* | *CI95%* |
|---|---|---|---|---|
| *Population density rate* | per km$^2$ | 37.4 [27.9; 46.0] | 1.1 | [0.72;1.69] |
| *Latitude* | - | 48.86 [48.84; 48.88] | 1.69 | [1.09;2.63] |
| *Longitude* | - | 2.35 [2.32; 2.38] | 3.01 | [1.9;4.91] |
| *0-2 years rate* | % | 3 [2;4] | 1.05 | [0.69;1.61] |
| *3-5 years rate* | % | 3 [2;3] | 1.53 | [1;2.38] |
| *6-10 years rate* | % | 4 [4;5] | 1.69 | [1.1;2.63] |
| *11-17 years rate* | % | 6 [5;8] | 1.61 | [1.05;2.5] |
| *18-24 years rate* | % | 10 [8;12] | 0.62 | [0.4;0.96] |
| *25-39 years rate* | % | 26 [20;31] | 0.49 | [0.31;0.76] |
| *40-54 years rate* | % | 20 [18;21] | 1 | [0.65;1.54] |
| *55-64 years rate* | % | 11 [10;12] | 1.78 | [1.15;2.77] |
| *65-79 years rate* | % | 12 [9;14] | 1.05 | [0.69;1.61] |
| *Above 80 years rate* | % | 5 [3;7] | 0.76 | [0.49;1.16] |
| *Men rate* | % | 47 [45;49] | 1.27 | [0.83;1.95] |
| *French nationality rate* | % | 87 [83;89] | 0.49 | [0.31;0.76] |
| *Foreigners rate* | % | 13 [11;17] | 2.06 | [1.33;3.26] |
| *Immigrants rate* | % | 19 [16;23] | 3.2 | [2.01;5.25] |
| *Family rate* | % | 100 [98;100] | 0.72 | [0.47;1.11] |

| *Indicators* | *Units* | *Median [IQR]* | *OR* | *CI95%* |
|---|---|---|---|---|
| *Above 15 years rate* | % | 86 [84;88] | 0.49 | [0.31;0.76] |
| *0-14 years rate* | % | 14 [12;16] | 1.96 | [1.27;3.08] |
| *15-29 years rate* | % | 23 [20;26] | 0.62 | [0.4;0.96] |
| *30-44 years rate* | % | 22 [19;26] | 0.72 | [0.47;1.11] |
| *45-59 years rate* | % | 18 [16;20] | 1.87 | [1.21;2.92] |
| *60-74 years rate* | % | 14 [12;16] | 1.21 | [0.79;1.86] |
| *Above 75 years rate* | % | 8 [5;10] | 0.87 | [0.57;1.33] |
| *0-19 years rate* | % | 19 [16;22] | 1.69 | [1.1;2.63] |
| *20-64 years rate* | % | 65 [60;69] | 0.57 | [0.36;0.87] |
| *Above 65 years rate* | % | 16 [13;20] | 0.96 | [0.62;1.47] |
| *Farmer operators rate* | % | 0 [0;0] | 0.44 | [0.17;0.96] |
| *Craftsmen, businessmen, shopkeepers rate* | % | 3 [3;4] | 0.33 | [0.2;0.53] |
| *Intermediate professions rate* | % | 31 [24;36] | 0.19 | [0.1;0.31] |
| *Unskilled workers rate* | % | 15 [12;17] | 1.21 | [0.79;1.86] |
| *Blue collar unskilled workers rate* | % | 11 [9;15] | 4.38 | [2.67;7.51] |
| *White collar unskilled workers rate* | % | 4 [2;6] | 3.61 | [2.24;6.01] |
| *Retirees rate* | % | 18 [15;22] | 1.27 | [0.83;1.95] |
| *Other activities rate* | % | 17 [14;20] | 1.61 | [1.05;2.5] |

| *Indicators for men* | *Units* | *Median [IQR]* | *OR* | *CI95%* |
|---|---|---|---|---|
| *Above 15 years rate* | % | 85 [83;88] | 0.57 | [0.36;0.87] |
| *0-14 years rate* | % | 15 [12;18] | 1.78 | [1.15;2.77] |
| *15-29 years rate* | % | 23 [20;26] | 0.72 | [0.47;1.11] |
| *30-44 years rate* | % | 23 [20;27] | 0.72 | [0.47;1.11] |
| *45-59 years rate* | % | 19 [16;20] | 1.05 | [0.69;1.61] |
| *60-74 years rate* | % | 13 [11;16] | 1.27 | [0.83;1.95] |
| *Above 75 years rate* | % | 6 [4;8] | 0.79 | [0.52;1.21] |
| *0-19 years rate* | % | 20 [17;24] | 1.78 | [1.15;2.77] |
| *20-64 years rate* | % | 65 [60;70] | 0.62 | [0.4;0.96] |
| *Above 65 years rate* | % | 14 [11;18] | 1.15 | [0.75;1.77] |
| *Farmer operators rate* | % | 0 [0;0] | 0.54 | [0.16;1.35] |
| *Craftsmen, businessmen, shopkeepers rate* | % | 5 [4;7] | 0.44 | [0.28;0.68] |
| *Intermediate professions rate* | % | 35 [28;40] | 0.23 | [0.13;0.38] |
| *Unskilled workers rate* | % | 13 [11;16] | 1.46 | [0.95;2.26] |
| *Blue collar unskilled workers rate* | % | 9 [6;11] | 4.1 | [2.52;6.96] |
| *White collar unskilled workers rate* | % | 6 [4;9] | 4.1 | [2.52;6.96] |
| *Retirees rate* | % | 17 [13;20] | 1.21 | [0.79;1.86] |
| *Other activities rate* | % | 14 [12;18] | 1.96 | [1.27;3.08] |

| *Indicators for women* | *Units* | *Median [IQR]* | *OR* | *CI95%* |
|---|---|---|---|---|
| *Above 15 years rate* | % | 87 [85;90] | 0.54 | [0.34;0.83] |
| *0-14 years rate* | % | 13 [10;15] | 1.69 | [1.1;2.63] |

| | | | | |
|---|---|---|---|---|
| *15-29 years rate* | % | 23 [20;27] | 0.51 | [0.33;0.79] |
| *30-44 years rate* | % | 21 [18;25] | 0.72 | [0.47;1.11] |
| *45-59 years rate* | % | 18 [16;20] | 2.17 | [1.4;3.44] |
| *60-74 years rate* | % | 15 [12;17] | 1 | [0.65;1.54] |
| *Above 75 years rate* | % | 9 [6;12] | 0.91 | [0.6;1.4] |
| *0-19 years rate* | % | 17 [15;20] | 1.69 | [1.1;2.63] |
| *20-64 years rate* | % | 64 [59;68] | 0.76 | [0.49;1.16] |
| *Above 65 years rate* | % | 18 [14;22] | 0.87 | [0.57;1.33] |
| *Craftsmen, businessmen, shopkeepers rate* | % | 2 [1;3] | 0.42 | [0.26;0.65] |
| *Intermediate professions rate* | % | 27 [21;32] | 0.19 | [0.1;0.31] |
| *Unskilled workers rate* | % | 16 [13;18] | 0.96 | [0.62;1.47] |
| *Blue collar unskilled workers rate* | % | 14 [11;18] | 3.84 | [2.37;6.46] |
| *White collar unskilled workers rate* | % | 2 [1;3] | 3.01 | [1.9;4.92] |
| *Retirees rate* | % | 20 [16;24] | 1.27 | [0.83;1.95] |
| *Other activities rate* | % | 18 [15;22] | 1.46 | [0.95;2.26] |

Housing indicators

| *Indicators* | *Units* | *Median [IQR]* | *OR* | *CI95%* |
|---|---|---|---|---|
| *Primary residences* | % | 84 [80;89] | 5.87 | [3.46;10.61] |
| *Secondary residences* | % | 7 [4;11] | 0.26 | [0.16;0.42] |
| *Vacant homes* | % | 8 [6;10] | 0.69 | [0.44;1.06] |
| *Detached houses* | % | 1 [0;1] | 0.91 | [0.6;1.4] |
| *Flats* | % | 98 [96;99] | 1.05 | [0.69;1.61] |
| *Single person housing* | % | 22 [17;28] | 0.79 | [0.52;1.21] |
| *Two persons housing* | % | 31 [25;37] | 1.1 | [0.72;1.69] |
| *Three mersons housing* | % | 23 [18;27] | 1.69 | [1.1;2.63] |
| *Four persons housing* | % | 12 [9;16] | 1 | [0.65;1.54] |
| *Five and more persons housing* | % | 7 [4;12] | 0.49 | [0.31;0.76] |
| *Number of rooms in primary residencies* | - | 2.5 [2.3;2.8] | 1.05 | [0.69;1.61] |
| *Detached house primary residences* | % | 1 [0;1] | 0.91 | [0.6;1.4] |
| *Flats primary residence* | % | 98 [96;99] | 1.15 | [0.75;1.77] |
| *Primary residences below 30 sqm* | % | 21 [15;28] | 0.66 | [0.42;1.01] |
| *Primary residences between 30 and 40 sqm* | % | 16 [12;21] | 1.21 | [0.79;1.86] |
| *Primary residences between 40 and 60 sqm* | % | 24 [19;29] | 2.06 | [1.33;3.26] |
| *Primary residences between 60 and 80 sqm* | % | 16 [12;21] | 1.53 | [1;2.38] |
| *Primary residences between 80 and 100 sqm* | % | 9 [6;12] | 1.1 | [0.72;1.69] |
| *Primary residences between 100 and 120 sqm* | % | 4 [2;7] | 0.35 | [0.22;0.56] |
| *Primary residences above 120 sqm* | % | 2 [1;6] | 0.31 | [0.19;0.5] |
| *Primary residences built before 2014* | % | 100 [100;100] | 0.96 | [0.62;1.47] |
| *Primary residences built before 1919* | % | 31 [13;55] | 0.3 | [0.18;0.47] |
| *Primary residences built between 1919 and 1945* | % | 15 [10;23] | 0.62 | [0.4;0.96] |
| *Primary residences built between 1946 and 1970* | % | 16 [10;24] | 1.61 | [1.05;2.5] |
| *Primary residences built between 1971 and 1990* | % | 14 [6;27] | 2.17 | [1.4;3.44] |
| *Primary residences built between 1991 and 2005* | % | 4 [2;8] | 1.15 | [0.75;1.77] |
| *Primary residences built between 2006 and 2014* | % | 1 [0;1] | 1.61 | [1.05;2.5] |
| *Occupancy since less than two years* | % | 15 [12;18] | 0.49 | [0.31;0.76] |

| | | | | |
|---|---|---|---|---|
| *Households between two to four years* | % | 23 [20;26] | 0.42 | [0.26;0.65] |
| *Households between five to nine years* | % | 17 [15;19] | 0.83 | [0.54;1.27] |
| *Households above 10 years* | % | 44 [39;49] | 2.41 | [1.55;3.86] |
| *Primary residence owner* | % | 36 [27;42] | 0.35 | [0.22;0.56] |
| *Primary residence tenants* | % | 59 [52;69] | 2.85 | [1.8;4.62] |
| *Primary residence social housing* | % | 8 [1;25] | 4.69 | [2.84;8.14] |
| *Primary residence rent free accomodation* | % | 5 [3;6] | 0.25 | [0.14;0.4] |
| *Number of persons in primary residence* | - | 1.9 [1.7;2] | 1.61 | [1.05;2.5] |
| *Primary residence occupancy duration* | years | 13 [11;14] | 1.53 | [1;2.38] |
| *Primary residence with bathroom* | % | 93 [91;95] | 1.1 | [0.72;1.69] |
| *Primary residence with central heating* | % | 38 [20;62] | 2.29 | [1.47;3.64] |
| *Primary residence with individual heating* | % | 23 [14;32] | 0.49 | [0.31;0.76] |
| *Primary residence with electric heating* | % | 35 [20;49] | 0.54 | [0.34;0.83] |
| *Primary residence with carpark* | % | 24 [16;36] | 1.27 | [0.83;1.95] |
| *Primary residence occupants with at least one car* | % | 34 [27;43] | 0.66 | [0.42;1.01] |
| *Primary residence occupants with one car* | % | 31 [24;37] | 0.66 | [0.42;1.01] |
| *Primary residence occupants with two car* | % | 3 [2;5] | 0.62 | [0.4;0.96] |
| *Mean number of persons in primary residence owners* | - | 1.8 [1.65;2] | 2.85 | [1.8;4.62] |
| *Primary residence owners' occupancy mean duration* | years | 8.8 [7.3;11.5] | 2.55 | [1.63;4.09] |
| *Mean number of persons in social housing primary residence* | - | 2.2 [1.8;2.6] | 2.19 | [1.4;3.5] |
| *Social housing primary residence occupancy mean duration* | years | 14 [10;18] | 1.25 | [0.81;1.94] |
| *Mean number of persons in rent free primary residence* | - | 1.7 [1.5;2] | 1.26 | [0.83;1.95] |
| *Rent free primary residence occupancy mean duration* | years | 10 [8;12] | 0.87 | [0.57;1.33] |

Education indicators

| *Indicators* | *Units* | *Median [IQR]* | *OR* | *CI95%* |
|---|---|---|---|---|
| *2-5 years children attending school rate among this age group* | % | 72 [65;78] | 1.26 | [0.82;1.94] |
| *6-10 years children attending school among this age group* | % | 99 [96;100] | 0.51 | [0.32;0.79] |
| *11-14 years children attending school among this age group* | % | 100 [98;100] | 0.5 | [0.33;0.77] |
| *15-17 years children attending school among this age group* | % | 100 [96;100] | 0.78 | [0.51;1.21] |
| *18-24 years adults attending school among this age group* | % | 73 [65;79] | 0.54 | [0.34;0.83] |
| *25-29 years adults attending school among this age group* | % | 16 [12;21] | 1.04 | [0.68;1.59] |
| *Over 30 years adults attending school among this age group* | % | 2 [2;3] | 1.32 | [0.86;2.04] |
| *No diploma rate among workforce over 15 years* | % | 15 [12;21] | 3.84 | [2.37;6.46] |
| *Vocational school graduates rate among workforce over 15 years* | % | 7 [5;10] | 4.38 | [2.67;7.51] |
| *High school graduates rate among workforce over 15 years* | % | 13 [11;15] | 2.69 | [1.71;4.34] |
| *High education graduates rate among workforce over 15 years* | % | 65 [54;71] | 0.2 | [0.11;0.33] |
| *No diploma men rate among workforce men over 15 years* | % | 14 [11;20] | 4.38 | [2.67;7.51] |
| *Vocational school graduates men rate among workforce men over 15 years* | % | 8 [6;11] | 4.69 | [2.84;8.14] |

| | | | | |
|---|---|---|---|---|
| *High school graduates men rate among workforce men over 15 years* | % | 12 [10;15] | 2.69 | [1.71;4.34] |
| *High education graduates men rate among workforce men over 15 years* | % | 66 [54;73] | 0.2 | [0.11;0.33] |
| *No diploma women rate among workforce women over 15 years* | % | 16 [13;22] | 5.04 | [3.03;8.85] |
| *Vocational school graduates women rate among workforce women over 15 years* | % | 7 [5;10] | 3.84 | [2.37;6.46] |
| *High school graduates women rate among workforce women over 15 years* | % | 13 [11;16] | 1.78 | [1.15;2.77] |
| *High education graduates women rate among workforce women over 15 years* | % | 64 [55;69] | 0.19 | [0.1;0.31] |

Family indicators

| *Indicators* | *Units* | *Median [IQR]* | *OR* | *CI95%* |
|---|---|---|---|---|
| *Single person household rate* | % | 51 [46;56] | 0.76 | [0.49;1.16] |
| *Single men household rate* | % | 21 [18;25] | 0.96 | [0.62;1.47] |
| *Single women household rate* | % | 30 [27;33] | 0.42 | [0.26;0.65] |
| *Other household without family rate* | % | 4 [3;6] | 1.1 | [0.72;1.69] |
| *Household with a family rate* | % | 44 [39;50] | 1.33 | [0.87;2.05] |
| *Household with a child-free family rate* | % | 19 [17;22] | 0.33 | [0.2;0.53] |
| *Houhold with a family with children rate* | % | 16 [13;21] | 1.05 | [0.69;1.61] |
| *Household with a single-parent family rate* | % | 7 [6;10] | 3.2 | [2.01;5.25] |
| *Rate of household with farmer operators household head* | % | 0 [0;0] | 0.36 | [0.11;0.89] |
| *Rate of household with craftsmen, businessmen, shopkeepers household head* | % | 4 [3;5] | 0.59 | [0.38;0.92] |
| *Rate of household with highly qualified occupation household head* | % | 35 [29;39] | 0.21 | [0.12;0.35] |
| *Rate of household with intermediate professions household head* | % | 16 [13;19] | 1.33 | [0.87;2.05] |
| *Rate of household with blue collar unskilled workers household head* | % | 12 [10;16] | 4.38 | [2.67;7.51] |
| *Rate of household with white collar unskilled workers household head* | % | 4 [3;6] | 3.39 | [2.12;5.61] |
| *Rate of household with retirees household head* | % | 20 [16;24] | 1.27 | [0.83;1.95] |
| *Rate of household with other activities household head* | % | 7 [6;9] | 0.62 | [0.4;0.96] |
| *Rate of persons among population over 15 years living in single person household* | % | 27 [23;32] | 0.66 | [0.42;1.01] |
| *Rate of persons among population over 15 years living in single men household* | % | 11 [9;14] | 0.76 | [0.49;1.16] |
| *Rate of persons among population over 15 years living in single women household* | % | 16 [13;19] | 0.44 | [0.28;0.68] |
| *Rate of persons among population over 15 years living in other household without family* | % | 5 [4;7] | 0.91 | [0.6;1.4] |
| *Rate of persons among population over 15 years living in household with a family* | % | 67 [61;72] | 1.27 | [0.83;1.95] |
| *Rate of persons among population over 15 years living in household with a child-free family* | % | 22 [18;24] | 0.33 | [0.2;0.53] |
| *Rate of persons among population over 15 years living in household with a family with children* | % | 34 [29;40] | 1.05 | [0.69;1.61] |

| | | | | |
|---|---|---|---|---|
| *Rate of persons among population over 15 years living in household with a single-parent family* | % | 10 [8;13] | 3.2 | [2.01;5.25] |
| *Rate of persons among population over 15 years living with a farmer operators household head* | % | 0 [0;0] | 0.36 | [0.11;0.89] |
| *Rate of persons among population over 15 years living with craftsmen, businessmen, shopkeepers household head* | % | 5 [4;7] | 0.54 | [0.34;0.83] |
| *Rate of persons among population over 15 years living with a highly qualified occupation household head* | % | 39 [31;44] | 0.21 | [0.12;0.35] |
| *Rate of persons among population over 15 years living with an intermediate professions household head* | % | 16 [14;19] | 1.61 | [1.05;2.5] |
| *Rate of persons among population over 15 years living with a blue collar unskilled workers household head* | % | 12 [10;18] | 4.38 | [2.67;7.51] |
| *Rate of persons among population over 15 years living with a white collar unskilled workers household head* | % | 5 [3;8] | 3.84 | [2.37;6.46] |
| *Rate of persons among population over 15 years livingwith a retirees household head* | % | 15 [12;18] | 1 | [0.65;1.54] |
| *Rate of persons among population over 15 years living with other activities household head* | % | 5 [4;7] | 1 | [0.65;1.54] |
| *15-24 years rate among population over 15 years* | % | 15 [13;17] | 0.96 | [0.62;1.47] |
| *25-54 years rate among population over 15 years* | % | 52 [47;58] | 0.83 | [0.54;1.27] |
| *55-79 years rate among population over 15 years* | % | 26 [23;30] | 1.78 | [1.15;2.77] |
| *over 80 years rate among population over 15 years* | % | 5 [4;8] | 0.76 | [0.49;1.16] |
| *Singles rate among population over 15 years* | % | 31 [26;36] | 0.66 | [0.42;1.01] |
| *Married persons rate among population over 15 years* | % | 32 [28;37] | 0.91 | [0.6;1.4] |
| *Non-married persons rate among population over 15 years* | % | 68 [63;72] | 1.1 | [0.72;1.69] |
| *15-24 years rate among household population over 15 years* | % | 15 [13;17] | 1.1 | [0.72;1.69] |
| *25-54 years rate among household population over 15 years* | % | 53 [47;58] | 0.79 | [0.52;1.21] |
| *55-79 years rate among household population over 15 years* | % | 26 [23;30] | 1.69 | [1.1;2.63] |
| *Over 80 years rate among household population over 15 years* | % | 5 [4;7] | 0.72 | [0.47;1.11] |
| *15-24 years rate among single person households* | % | 10 [7;15] | 0.37 | [0.23;0.59] |
| *25-54 years rate among single person households* | % | 47 [40;55] | 0.72 | [0.47;1.11] |
| *55-79 years rate among single person households* | % | 30 [25;36] | 3.2 | [2.01;5.25] |
| *Over 80 years rate among single person households* | % | 9 [6;14] | 1 | [0.65;1.54] |
| *Family with children rate among family households* | % | 37 [33;42] | 1.1 | [0.72;1.69] |
| *Single parent family rate among family households* | % | 17 [13;22] | 3.84 | [2.37;6.46] |
| *Family without children rate among family households* | % | 45 [38;51] | 0.33 | [0.2;0.53] |
| *Family without children below 25 years rate among family households* | % | 51 [45;56] | 0.39 | [0.25;0.62] |
| *Family with one child below 25 years rate among family households* | % | 24 [21;26] | 1.96 | [1.27;3.08] |
| *Family with two children below 25 years rate among family households* | % | 17 [15;20] | 1.16 | [0.76;1.78] |
| *Family with three children below 25 years rate among family households* | % | 6 [4;8] | 2.55 | [1.63;4.09] |
| *Family with more than 4 children below 25 years rate among family households* | % | 2 [1;3] | 2.41 | [1.55;3.86] |

Employment indicators

| Indicators | Units | Median [IQR] | OR | CI95% |
|---|---|---|---|---|
| *15-64 years workforce rate among population of the same age group* | % | 79 [74;82] | 0.49 | [0.31;0.76] |
| *15-24 years workforce rate among population of the same age group* | % | 36 [30;43] | 1.27 | [0.83;1.95] |
| *25-54 years workforce rate among population of the same age group* | % | 92 [90;94] | 0.42 | [0.26;0.65] |
| *55-64 years workforce rate among population of the same age group* | % | 72 [67;77] | 0.37 | [0.23;0.59] |
| *15-64 years men workforce rate among men of the same age group* | % | 81 [77;85] | 0.46 | [0.29;0.72] |
| *15-24 years men workforce rate among men of the same age group* | % | 36 [29;44] | 1.33 | [0.87;2.05] |
| *25-54 years men workforce rate among men of the same age group* | % | 94 [93;96] | 0.42 | [0.26;0.65] |
| *55-64 years men workforce rate among men of the same age group* | % | 77 [70;83] | 0.36 | [0.22;0.56] |
| *15-64 years women workforce rate among women of the same age group* | % | 76 [72;80] | 0.76 | [0.49;1.16] |
| *15-24 years women workforce rate among women of the same age group* | % | 37 [29;43] | 1.33 | [0.87;2.05] |
| *25-54 years women workforce rate among women of the same age group* | - | 91 [88;93] | 0.35 | [0.22;0.56] |
| *55-64 years women workforce rate among women of the same age group* | % | 68 [63;74] | 0.57 | [0.36;0.87] |
| *15-64 years employed workforce rate among population workforce of the same age group* | % | 89 [86;91] | 0.3 | [0.18;0.47] |
| *15-24 years employed workforce rate among population workforce of the same age group* | % | 81 [74;87] | 0.21 | [0.12;0.35] |
| *25-54 years employed workforce rate among population workforce of the same age group* | % | 89 [87;91] | 0.33 | [0.2;0.53] |
| *55-64 years employed workforce rate among population workforce of the same age group* | % | 89 [85;93] | 0.49 | [0.31;0.76] |
| *15-64 years men employed workforce among men workforce of the same age group* | % | 89 [86;91] | 0.28 | [0.17;0.45] |
| *15-24 years men employed workforce among men workforce of the same age group* | % | 80 [70;89] | 0.36 | [0.22;0.57] |
| *25-54 years men employed workforce among men workforce of the same age group* | % | 90 [87;92] | 0.35 | [0.22;0.56] |
| *55-64 years men employed workforce among men workforce of the same age group* | % | 89 [83;94] | 0.59 | [0.38;0.92] |
| *15-64 years women employed workforce among women workforce of the same age group* | % | 88 [86;90] | 0.26 | [0.16;0.42] |
| *15-24 years women employed workforce among women workforce of the same age group* | % | 82 [74;89] | 0.33 | [0.2;0.53] |
| *25-54 years women employed workforce among women workforce of the same age group* | % | 89 [86;91] | 0.4 | [0.25;0.62] |
| *55-64 years women employed workforce among women workforce of the same age group* | % | 90 [86;94] | 0.37 | [0.23;0.59] |
| *15-64 years unemployed workforce among population workforce of the same age group* | % | 13 [10;16] | 3.39 | [2.12;5.61] |
| *15-24 years unemployed workforce among population workforce of the same age group* | % | 23 [15;35] | 4.69 | [2.84;8.14] |
| *25-54 years unemployed workforce among population workforce of the same age group* | % | 12 [9;15] | 3.01 | [1.9;4.92] |

| | | | | |
|---|---|---|---|---|
| *55-64 years unemployed workforce among population workforce of the same age group* | % | 12 [8;18] | 1.96 | [1.27;3.08] |
| *15-64 years non-working population among population of the same age group* | % | 8 [7;10] | 1.96 | [1.27;3.08] |
| *15-64 years studying population among non-working population of the same age group* | % | 27 [22;34] | 0.87 | [0.57;1.33] |
| *15-64 years retired population among non-working population of the same age group* | % | 15 [12;20] | 2.55 | [1.63;4.09] |
| *15-64 years other inactive population among non-working population of the same age group* | % | 4 [3;5] | 3.61 | [2.24;6.01] |
| *15-64 years unemployed men among men workforce of the same age group* | % | 7 [5;10] | 3.61 | [2.24;6.01] |
| *15-64 years non-working population among population of the same age group* | % | 11 [9;14] | 2.17 | [1.4;3.44] |
| *15-64 years studying population among non-working population of the same age group* | % | 23 [18;30] | 1 | [0.65;1.54] |
| *15-64 years retired population among non-working population of the same age group* | % | 14 [11;19] | 2.69 | [1.71;4.34] |
| *15-64 years other inactive population among non-working population of the same age group* | % | 3 [2;4] | 3.39 | [2.12;5.61] |
| *15-64 years unemployed women among women workforce of the same age group* | % | 5 [3;7] | 4.1 | [2.52;6.96] |
| *15-64 years non-working population among population of the same age group* | % | 12 [10;14] | 1.27 | [0.83;1.95] |
| *15-64 years studying population among non-working population of the same age group* | % | 31 [25;39] | 0.87 | [0.57;1.33] |
| *15-64 years retired population among non-working population of the same age group* | % | 16 [13;21] | 1.7 | [1.1;2.65] |
| *15-64 years other inactive population among non-working population of the same age group* | % | 4 [3;5] | 2.69 | [1.71;4.34] |
| *Farmer operators rate among 15-64 years population workforce* | % | 9 [7;13] | 0.52 | [0.2;1.13] |
| *Craftsmen, businessmen, shopkeepers rate among 15-64 years population workforce* | % | 0 [0;0] | 0.39 | [0.25;0.62] |
| *Highly qualified worker among 15-64 years population workforce* | % | 5 [4;6] | 0.23 | [0.13;0.38] |
| *Intermediate professions rate among 15-64 years population workforce* | % | 48 [38;53] | 1.46 | [0.95;2.26] |
| *Blue collar unskilled workers rate among 15-64 years population workforce* | % | 23 [20;25] | 5.04 | [3.03;8.85] |
| *White collar unskilled workers rate among 15-64 years population workforce* | % | 17 [14;23] | 3.61 | [2.24;6.01] |
| *Farmer operators rate among 15-64 years population employed workforce* | % | 6 [4;8] | 0.58 | [0.22;1.28] |
| *Craftsmen, businessmen, shopkeepers rate among 15-64 years population employed workforce* | % | 0 [0;0] | 0.44 | [0.28;0.68] |
| *Highly qualified worker among 15-64 years population employed workforce* | % | 5 [4;7] | 0.23 | [0.13;0.38] |
| *Intermediate professions rate among 15-64 years population employed workforce* | % | 51 [40;56] | 1.69 | [1.1;2.63] |
| *Blue collar unskilled workers rate among 15-64 years population employed workforce* | % | 22 [19;25] | 4.38 | [2.67;7.51] |
| *White collar unskilled workers rate among 15-64 years population employed workforce* | % | 16 [13;21] | 4.1 | [2.52;6.96] |
| *Men rate among employed workforce over 15 years* | % | 5 [3;7] | 1.2 | [0.79;1.85] |
| *Women rate among employed workforce over 15 years* | % | 49 [47;51] | 0.87 | [0.56;1.33] |

| | | | | |
|---|---|---|---|---|
| *Salaried employees rate among employed workforce over 15 years* | % | 51 [49;53] | 3.37 | [2.11;5.58] |
| *Non-salaried employees rate among employed workforce over 15 years* | % | 84 [79;88] | 0.29 | [0.18;0.47] |
| *Full-time employees rate among employed workforce over 15 years* | % | 16 [12;21] | 2.05 | [1.32;3.24] |
| *Working in Paris rate among employed workforce over 15 years* | % | 16 [14;18] | 1.39 | [0.9;2.14] |
| *Working outside Paris rate among employed workforce over 15 years* | % | 69 [65;72] | 0.72 | [0.46;1.1] |
| *Working in Ile-de-France region except Paris rate among employed workforce over 15 years* | % | 31 [28;35] | 0.72 | [0.46;1.1] |
| *Working in France except Ile-de-France region rate among employed workforce over 15 years* | % | 30 [26;33] | 0.87 | [0.56;1.33] |
| *Working outside France rate among employed workforce over 15 years* | % | 1 [1;2] | 0.41 | [0.26;0.65] |
| *Not using any means of transport to work rate among employed workforce over 15 years* | % | 0 [0;1] | 0.46 | [0.29;0.72] |
| *Walking to work rate among employed workforce over 15 years* | % | 100 [100;100] | 0.51 | [0.32;0.79] |
| *Two-wheelers to work rate among employed workforce over 15 years* | % | 4 [3;6] | 0.41 | [0.26;0.65] |
| *Driving to work rate among employed workforce over 15 years* | % | 9 [7;12] | 0.75 | [0.49;1.15] |
| *Using public transportation to work rate among employed workforce over 15 years* | % | 9 [7;11] | 3.18 | [2;5.22] |
| *Men rate among salaried workforce over 15 years* | % | 10 [8;14] | 1.61 | [1.05;2.5] |
| *Women rate among salaried workforce over 15 years* | % | 66 [59;70] | 0.62 | [0.4;0.96] |
| *Men rate among non-salaried workforce over 15 years* | % | 47 [45;50] | 1.53 | [1;2.38] |
| *Women rate among non-salaried workforce over 15 years* | % | 53 [50;55] | 0.66 | [0.42;1.01] |
| *Men rate among full-time salaried workforce over 15 years* | % | 58 [54;63] | 1.33 | [0.87;2.05] |
| *Women rate among full-time salaried workforce over 15 years* | % | 42 [37;46] | 0.76 | [0.49;1.17] |
| *Salaried rate among full-time employed workforce over 15 years* | % | 32 [27;38] | 3.61 | [2.24;6.01] |
| *Non-salaried rate among full-time employed workforce over 15 years* | % | 68 [62;73] | 0.28 | [0.17;0.45] |
| *Full-time rate among salaried workforce over 15 years* | % | 79 [72;84] | 2.55 | [1.63;4.09] |
| *Permanent contracts rate among salaried workforce over 15 years* | % | 21 [16;28] | 0.62 | [0.4;0.96] |
| *Short-term contracts rate among salaried workforce over 15 years* | % | 15 [13;18] | 1.39 | [0.91;2.15] |
| *Interim contracts rate among salaried workforce over 15 years* | % | 84 [81;86] | 1.96 | [1.27;3.08] |
| *Subsidized work rate among salaried workforce over 15 years* | % | 11 [9;13] | 1.96 | [1.27;3.08] |
| *Apprenticeship contract rate among salaried workforce over 15 years* | % | 1 [0;1] | 0.62 | [0.4;0.96] |
| *Self-employed among non-salaried workforce over 15 years* | % | 0 [0;1] | 1.46 | [0.95;2.26] |
| *Employers among non-salaried workforce over 15 years* | % | 4 [3;5] | 0.51 | [0.33;0.79] |
| *Caregivers among non-salaried workforce over 15 years* | % | 66 [58;73] | 1.27 | [0.83;1.95] |
